# Biomedical Discovery through the integrative Biomedical Knowledge Hub (iBKH)

**DOI:** 10.1101/2021.03.12.21253461

**Authors:** Chang Su, Yu Hou, Suraj Rajendran, Jacqueline R. M. A. Maasch, Zehra Abedi, Haotan Zhang, Zilong Bai, Anthony Cuturrufo, Winston Guo, Fayzan F. Chaudhry, Gregory Ghahramani, Jian Tang, Feixiong Cheng, Yue Li, Rui Zhang, Jiang Bian, Fei Wang

**Author notes:** **Corresponding Author:** Fei Wang, 425 E 61 St. New York City. NY 10065. USA. Equal Contribution (Chang Su, Yu Hou).

## Abstract

The massive and continuously increasing volume of biomedical knowledge derived from biological experiments or gained from healthcare practices has become an invaluable treasure for biomedicine. The emerging biomedical knowledge graphs (BKGs) provide an efficient and effective way to manage the abundant knowledge in biomedical and life science. In the present study, we harmonized and integrated data from diverse biomedical resources to curate a comprehensive BKG, named the integrative Biomedical Knowledge Hub (iBKH). To facilitate the usage of iBKH in biomedical research, we developed a web-based, easy-to-use, publicly available graphical portal that allows fast, interactive, and visualized knowledge retrieval in iBKH. Furthermore, an efficient and scalable graph learning pipeline was developed for novel knowledge discovery in iBKH. As a proof of concept, we performed our iBKH-based method for computational in silico drug repurposing for Alzheimer’s disease. The iBKH is publicly available at: http://ibkh.ai/.

## Introduction

Biomedicine is a discipline with enormous volume of highly specialized biomedical knowledge accumulated from biological experiments and healthcare practices. In the past decades, efforts have been drawn to collect and manage the abundant biomedical knowledge and have resulted in diverse biomedical knowledge sources. For example, the biomedical ontologies (Rubin et al., 2008; Smith et al., 2005) store hierarchical relationship-based descriptions for biomedical entities, and the manually curated biomedical knowledge bases (Callahan et al., 2020; Zhu et al., 2019) store biomedical relational data. However, each knowledge source typically focuses on a sub-domain in biomedicine, and hence cannot provide a comprehensive perspective of life science. This hinders the efficient usage of cross-domain biomedical knowledge to provide system-level understanding of human diseases.

At this point, the biomedical knowledge graph (BKG) has become a novel paradigm for better management of the massive volume, sophisticated biomedical knowledge and has attracted significant attentions in recent years(Himmelstein et al., 2017; Nelson et al., 2019; Nicholson and Greene, 2020; Rotmensch et al., 2017; Santos et al., 2022; Sügis et al., 2019). Typically, a BKG is a graph or network that integrates, harmonizes, and stores biomedical knowledge collected from single or multiple expert-derived knowledge sources, where nodes are a set of biomedical entities (e.g., diseases, drugs, genes, biological processes, etc.) and edges between nodes/entities are relations linking the biomedical entities (e.g., drug-treats-disease, disease-associates-gene, drug-interacts-drug, etc.).(Himmelstein *et al*., 2017; Nicholson and Greene, 2020; Santos *et al*., 2022; Zhu et al., 2020)

In this context, although efforts have been made to construct BKGs by integrating diverse expert curated knowledge bases (Himmelstein *et al*., 2017; Li et al., 2020; Santos *et al*., 2022; Yu et al., 2019; Zhu *et al*., 2020) or by extracting knowledge from literature using natural language processing (NLP) techniques (Ernst et al., 2015; Percha and Altman, 2018; Yuan et al., 2020), they are not perfect and there remains the space to build a more comprehensive BKG to support advanced biomedical research. In addition, though these BKGs are publicly available, there remains a need for an open, easy-to-use user interface (UI) to fill the gap between the BKG and biomedical researchers and healthcare providers. To this end, this present study proposed a comprehensive BKG, termed the integrative Biomedical Knowledge Hub (iBKH), which was curated by integrating data from 18 high-quality and well-known knowledge sources, including biomedical ontologies, manually curated biomedical knowledge bases, existing BKGs, and NLP-extracted biomedical knowledge sources. To further demonstrate the use of iBKH in biomedical research, we developed a web-based, easy-to-use, intelligent graphical portal that allows fast, interactive knowledge retrieval in iBKH and visualization of the retrieved knowledge.

In addition, we introduced advanced graph learning approaches to the iBKH for computational knowledge discovery. Current graph learning techniques (Mohamed et al., 2021; Su et al., 2020), an emerging branch of machine learning and deep learning that can learn underlying knowledge from graph structure data, have advanced the application of BKG in accelerating novel biomedical knowledge discovery such as drug repurposing (Su et al., 2022; Zhang et al., 2021; Zhou et al., 2020; Zhu *et al*., 2020) and disease risk gene prioritization(Hu et al., 2021; Peng et al., 2021). In this context, we introduced the advanced graph learning approaches to the iBKH for computational knowledge discovery. Specifically, we designed a knowledge discovery module based on the DGL-KE (Deep Graph Library - Knowledge Embedding) software (Zheng et al., 2020) for efficient and scalable graph learning in iBKH. As a proof of concept, we demonstrated a use case of iBKH, armed with the graph learning algorithms, for in silico hypothesis generation for Alzheimer’s disease (AD) drug repurposing – one of the grand challenges in current biomedical research.

## Results

**Figure 1** illustrates overall pipeline of the present study, which includes the following modules including: 1) iBKH construction through biomedical knowledge integration, 2) development of graphical portal for fast knowledge retrieval based on iBKH, and 3) iBKH-based computational knowledge discovery through deep graph learning. **Figure 2** illustrates the schema of our BKG, i.e., iBKH. The iBKH is publicly available at: http://ibkh.ai/.

**Figure 1.**
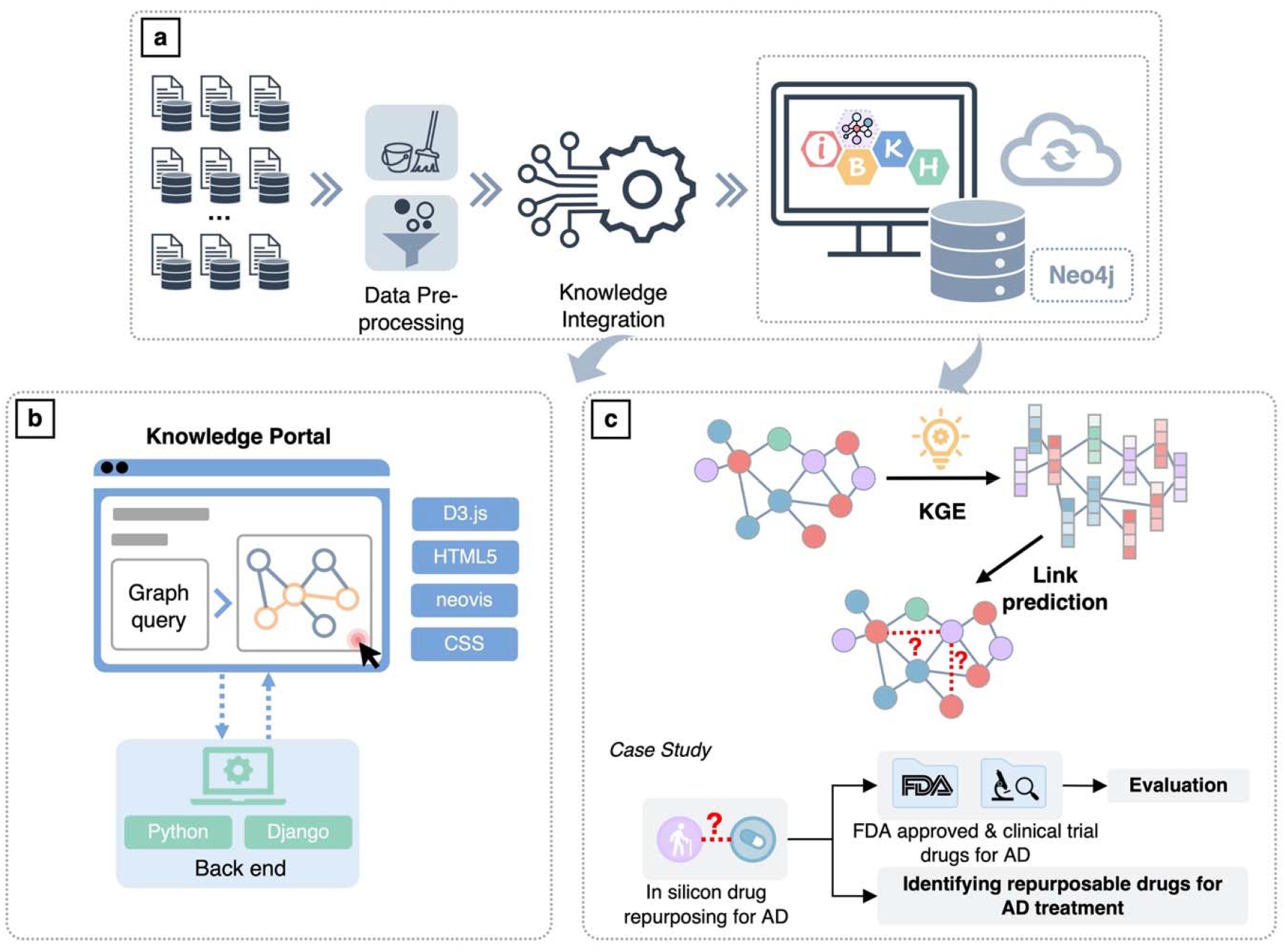
An illustration of study pipeline. **a**. Steps for curating iBKH. We first collected data from diverse biomedical data sources. Next, necessary data pre-processing, such as data cleaning and data filtering were performed. After that, knowledge from diverse sources were integrated to build an integrative knowledge graph, i.e., iBKH, which was deployed using Neo4j graph database. **b**. A web-based, easy-to-use graphical portal was developed for fast knowledge retrieval. **c**. A graph learning module was introduced to iBKH for novel knowledge discovery. Specifically, knowledge graph embedding was conducted to learn compressed vector representations for entities and relations in iBKH, which were further used for link prediction. As a proof of concept, we performed in silicon drug repurposing for Alzheimer’s disease. Abbreviations: AD = Alzheimer’s disease; CSS = Cascading Style Sheets; HTML5 = HyperText Markup Language Version 5; iBKH = integrative Biomedical Knowledge Hub; KGE = knowledge graph embedding.

**Figure 2.**
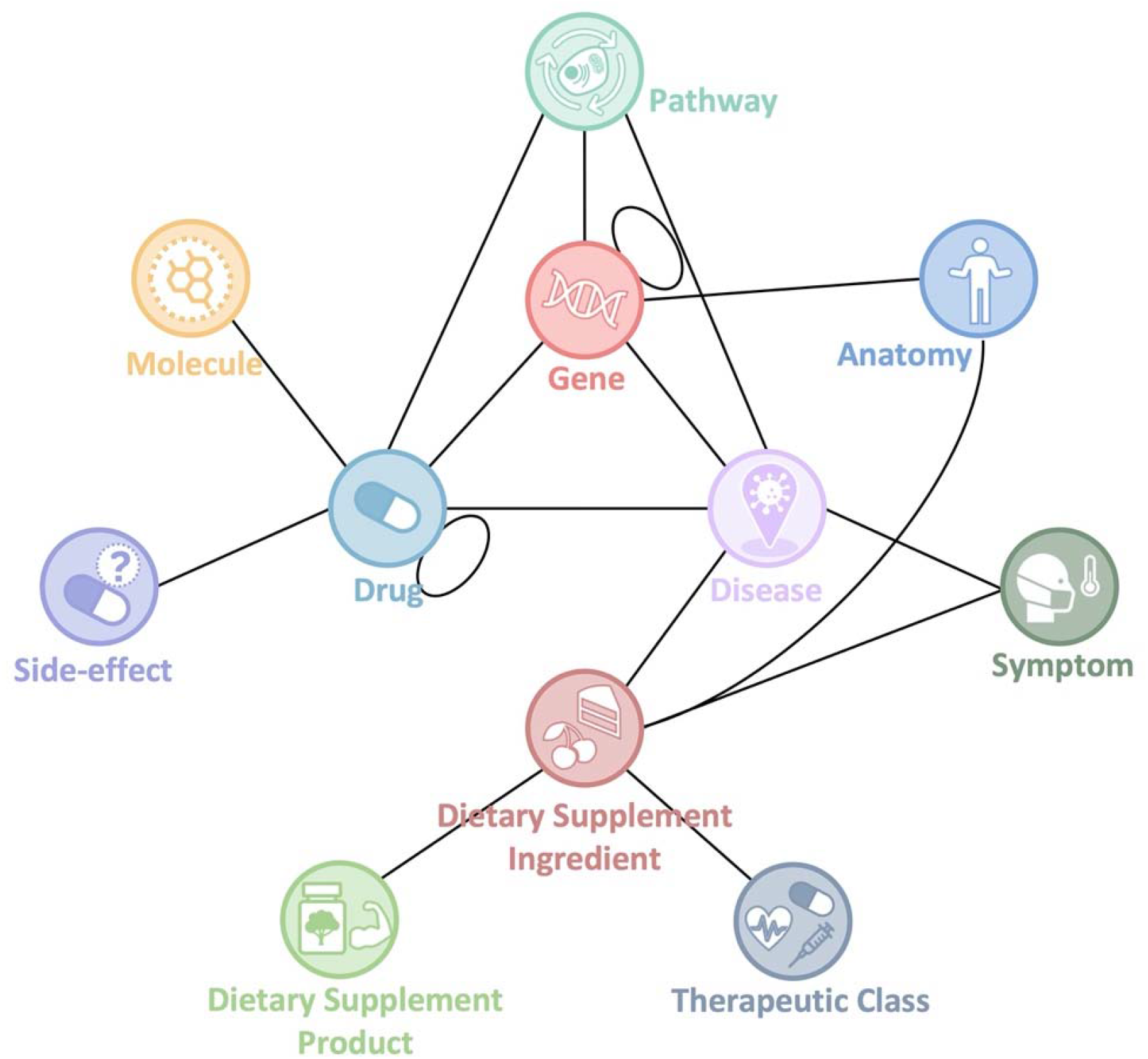
Schema of iBKH. Each circle denotes an entity type, and each link denotes a meta relation between a pair of entities. Of note, a meta relation can represent multiple types of relations between a specific pair of entities. For example, five potential relations including ‘Associates’, ‘Downregulates’, ‘Upregulates’, ‘Inferred_Relation’, ‘Text_Semantic_Relation’ can exist between a pair of disease and gene entities.

### The integrative Biomedical Knowledge Hub (iBKH)

By collecting, harmonizing, and integrating data from 18 publicly available biomedical knowledge sources (see **Table 1**), we curated a comprehensive biomedical knowledge graph, named the integrative Biomedical Knowledge Hub (iBKH). The knowledge sources include biomedical ontologies such as the Brenda Tissue Ontology (Chang et al., 2021), the Cell Ontology (Diehl et al., 2016) the Disease Ontology (Schriml et al., 2012), and the Uberon (Mungall et al., 2012); manually curated biomedical knowledge bases for biomedical entity and relation data such as the Bgee (Bastian et al., 2021), the Comparative Toxicogenomics Database (CTD) (Davis et al., 2019), the DrugBank,(Wishart et al., 2018) the Kyoto Encyclopedia of Genes and Genomes (KEGG) (Kanehisa and Goto, 2000), the Pharmacogenetics Knowledge Base (PharmGKB) (Hewett et al., 2002), the Reactome (Fabregat et al., 2018), the Side effect resource (SIDER)(Kuhn et al., 2016), and the TISSUE (Palasca et al., 2018); existing BKGs curated by integrating multiple knowledge bases such as the Drug Repurposing Knowledge Graph (DRKG, https://github.com/gnn4dr/DRKG) (Ioannidis et al., 2020), the Hetionet (Himmelstein *et al*., 2017), the Integrated Dietary Supplement Knowledge Base (iDISK) (Rizvi et al., 2020), our curated knowledge graph that covers a variety of dietary supplements, including vitamins, herbs, minerals, etc.; and other biomedical sources such as HUGO Gene Nomenclature Committee (HGNC) (Braschi et al., 2019), ChEMBL(Gaulton et al., 2012), and Chemical Entities of Biological Interest (ChEBI) (de Matos et al., 2010). More details of the sources can be found in **Table 1**.

**Table 1.**
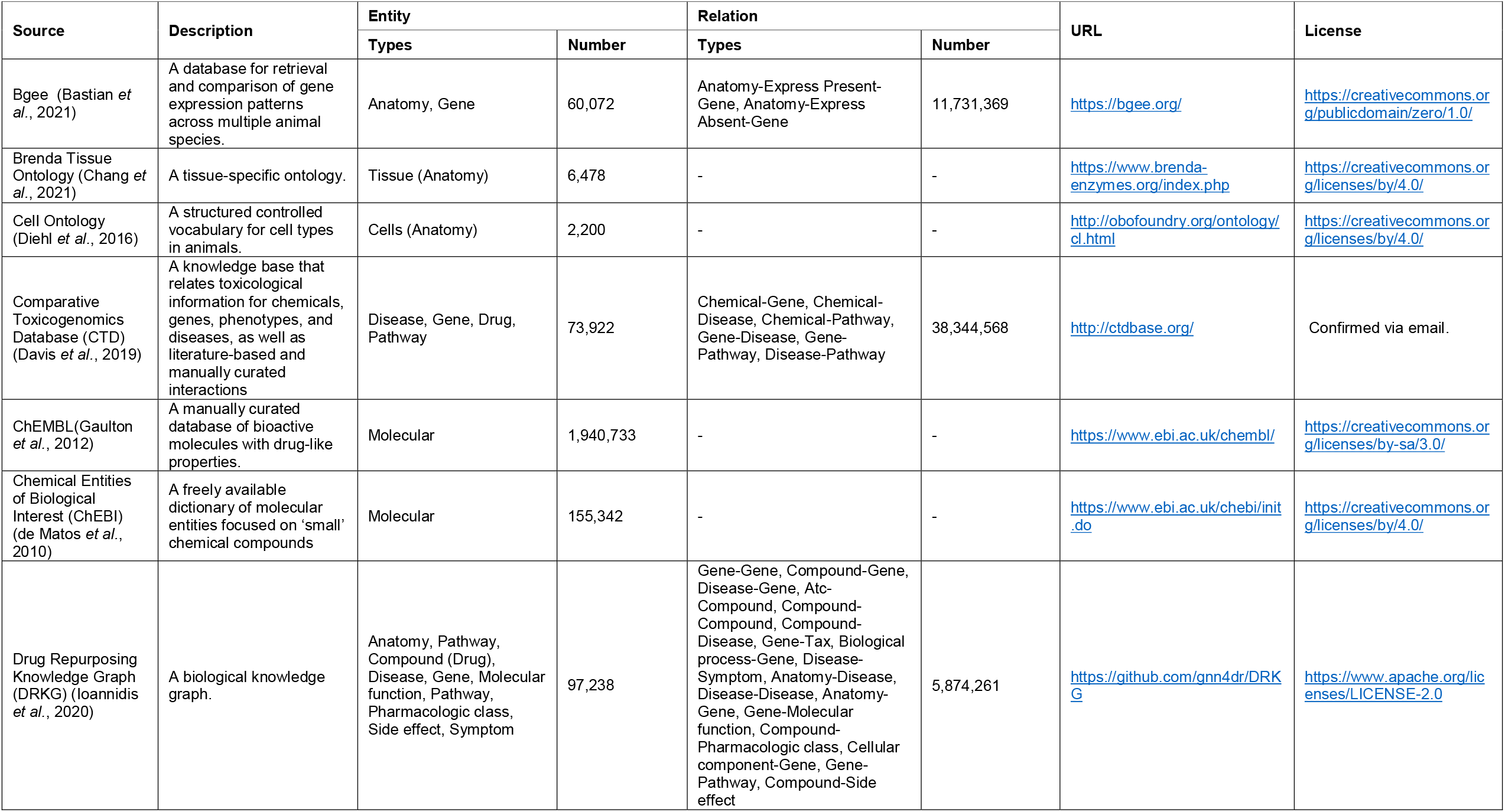
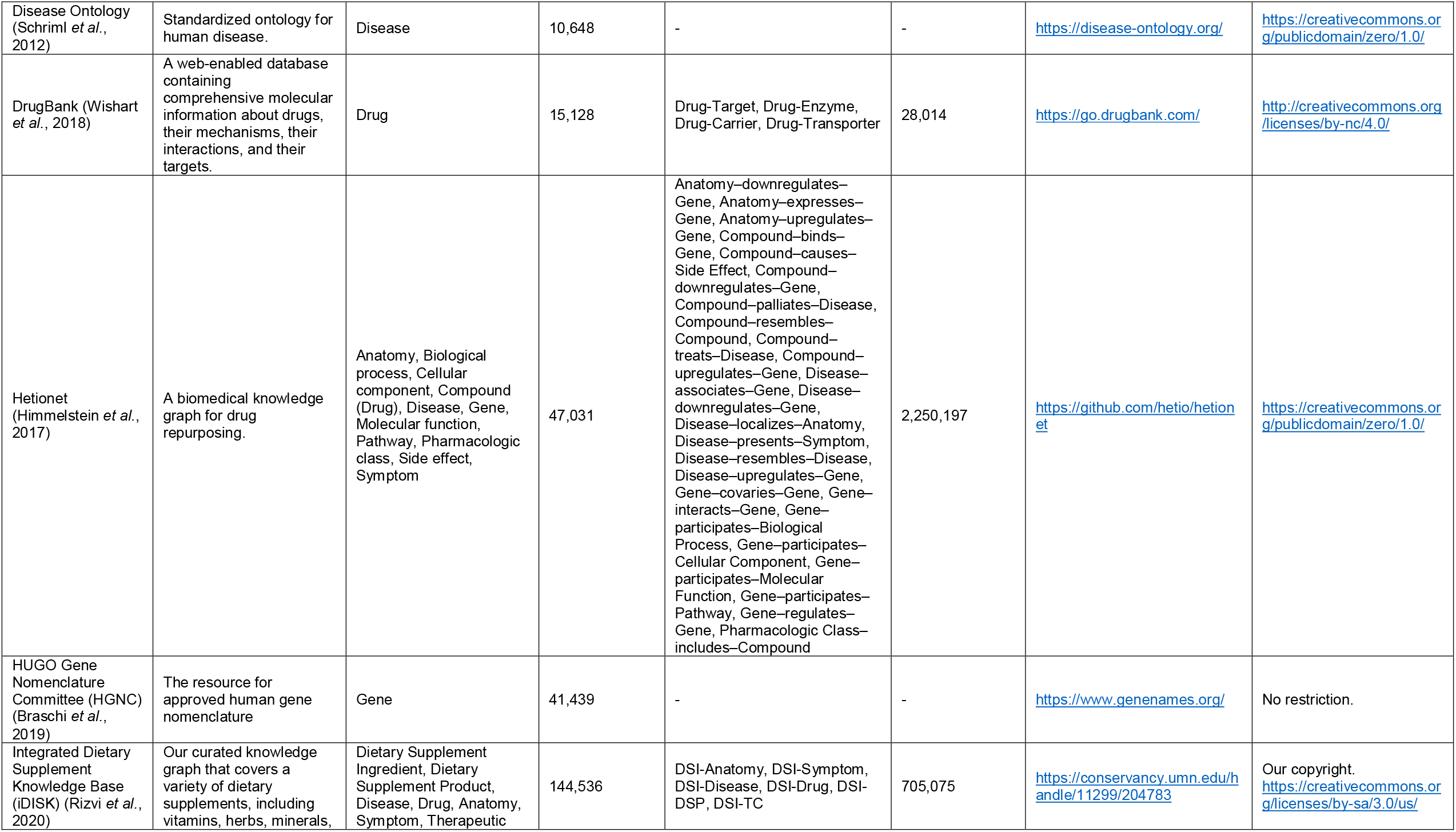
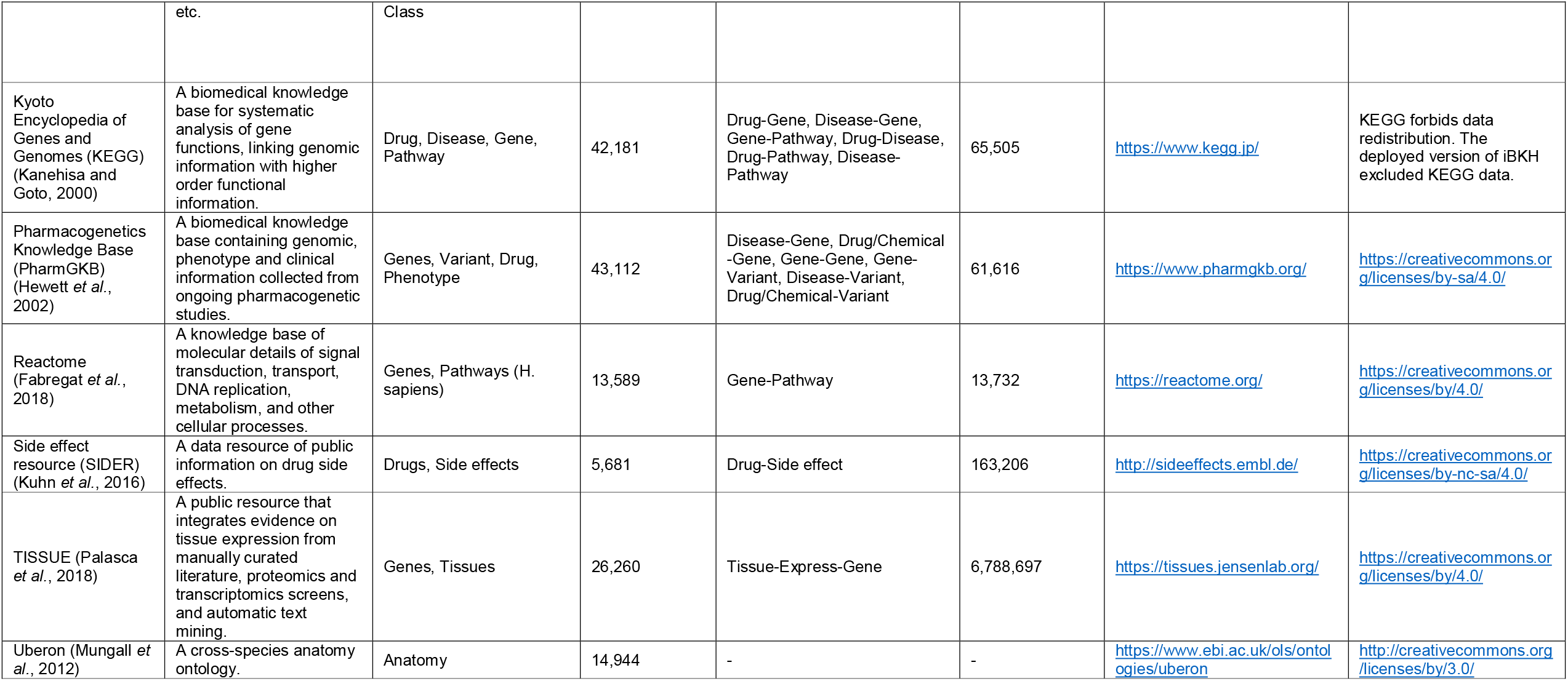
Data sources integrated for constructing iBKH.

After data management and necessary data cleaning, we integrated data from the diverse sources through biomedical entity term normalization and knowledge integration (more details can be found in the **Method** section). Current version of the resulted iBKH contains a total of 2,384,501 entities of 11 types, including 23,003 anatomy entities, 19,236 disease entities, 37,997 drug entities, 88,376 gene entities (including human and other species), 2,065,015 molecule entities, 1,361 symptom entities, 2,988 pathway entities, 4,251 side-effect entities, 4,101 dietary supplement ingredient (DSI) entities, 137,568 dietary supplement product (DSP) entities, 605 dietary’s therapeutic class (TC) entities (see **Figure 2** and **Table 2**). In addition, there are 45 relation types within 18 kinds of entity pairs, including Anatomy-Gene, Drug-Disease, Drug-Drug, Drug-Gene, Disease-Disease, Disease-Gene, Disease-Symptom, Gene-Gene, DSI-Disease, DSI-Symptom, DSI-Drug, DSI-Anatomy, DSI-DSP, DSI-TC, Disease-Pathway, Drug-Pathway, Gene-Pathway and Drug-Side Effect, which means multiple types of relations can exist between a pair of biomedical entities (see **Table 3**). Specifically, 2 types of potential relations can exist between a Anatomy-Gene pair, including ‘Expresses’ and ‘Absent’; 6 relation types between a Drug-Disease pair, such as ‘Treats’ and ‘Effects’; 2 relation types between a Drug-Drug pair including ‘Interaction’ and ‘Resembles’; 10 relation types between a Drug-Gene pair, such as ‘Targets’, ‘Upregulates’, and ‘Downregulates’; 2 relation types between a Disease-Disease pair including ‘Is_A’ and ‘Resembles’; 5 relation types between a Disease-Gene pair, such as ‘Associates’, ‘Upregulates’, and ‘Downregulates’; the ‘Presents’ relation type between a Disease-Symptom pair; and 5 relation types between a Gene-Gene pair, such as ‘Covaries’, ‘Interacts’, and ‘Regulates’; the ‘Has_Adverse Reaction’ relation between a DSI-Symptom pair; the ‘Is_Effective_For’ relation type between a DSI-Disease pair; the ‘Interacts’ relation type between a DSI-Drug pair; the ‘Has_Adverse_Effect_On’ relation type between a DSI-Anatomy pair; the ‘Has_Ingredient’ relation type between a DSI-DSP pair; the ‘Has_Therapeutic_Class’ relation type between a DSI-TC pair; the ‘Reaction’ and ‘Associates’ relation types between a Gene-Pathway pair; the ‘Associates’ relation between a Disease-Pathway pair; the ‘Associates’ relation between a Drug-pathway pair; the ‘Causes’ relation type between Drug-Side Effect pair.

**Table 2.**
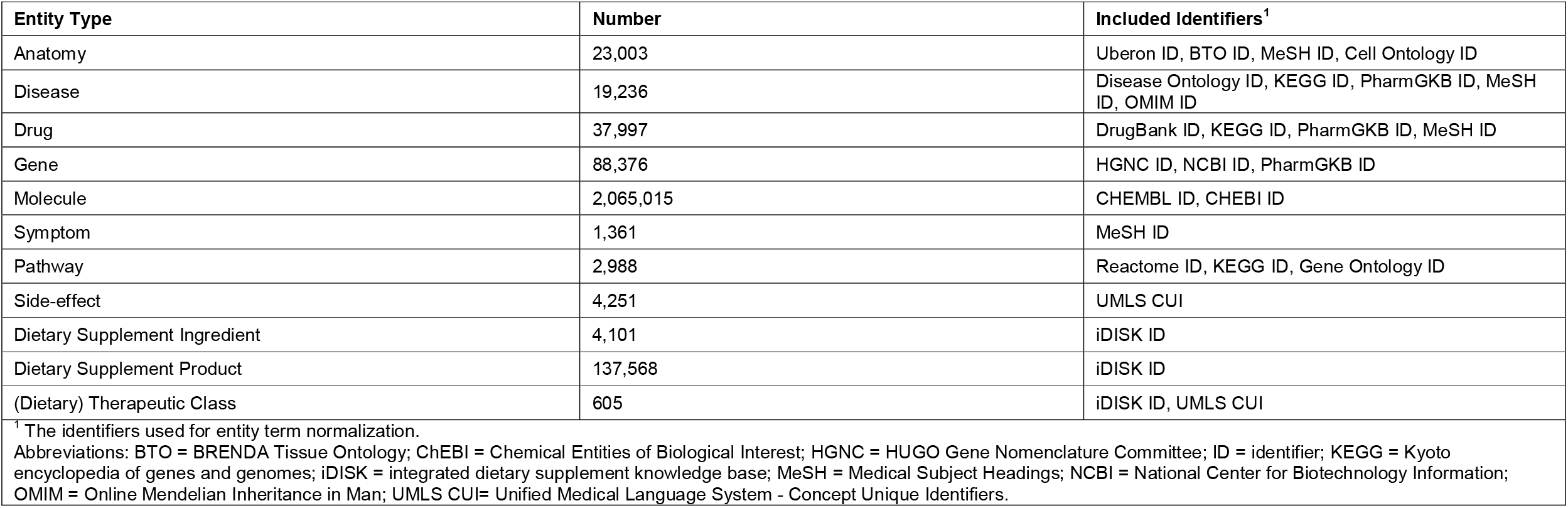
Statistics of biomedical entities in iBKH.

| Entity Type | Number | Included Identifiers<sup>1</sup> |
| --- | --- | --- |
| Anatomy | 23,003 | Uberon ID, BTO ID, MeSH ID, Cell Ontology ID |
| Disease | 19,236 | Disease Ontology ID, KEGG ID, PharmGKB ID, MeSH ID, OMIM ID |
| Drug | 37,997 | DrugBank ID, KEGG ID, PharmGKB ID, MeSH ID |
| Gene | 88,376 | HGNC ID, NCBI ID, PharmGKB ID |
| Molecule | 2,065,015 | CHEMBL ID, CHEBI ID |
| Symptom | 1,361 | MeSH ID |
| Pathway | 2,988 | Reactome ID, KEGG ID, Gene Ontology ID |
| Side-effect | 4,251 | UMLS CUI |
| Dietary Supplement Ingredient | 4,101 | iDISK ID |
| Dietary Supplement Product | 137,568 | iDISK ID |
| (Dietary) Therapeutic Class | 605 | iDISK ID, UMLS CUI |
| <sup>1</sup> The identifiers used for entity term normalization.<br>Abbreviations: BTO = BRENDA Tissue Ontology; ChEBI = Chemical Entities of Biological Interest; HGNC = HUGO Gene Nomenclature Committee; ID = identifier; KEGG = Kyoto encyclopedia of genes and genomes; iDISK = integrated dietary supplement knowledge base; MeSH = Medical Subject Headings; NCBI = National Center for Biotechnology Information; OMIM = Online Mendelian Inheritance in Man; UMLS CUI= Unified Medical Language System - Concept Unique Identifiers. |  |  |

**Table 3.**
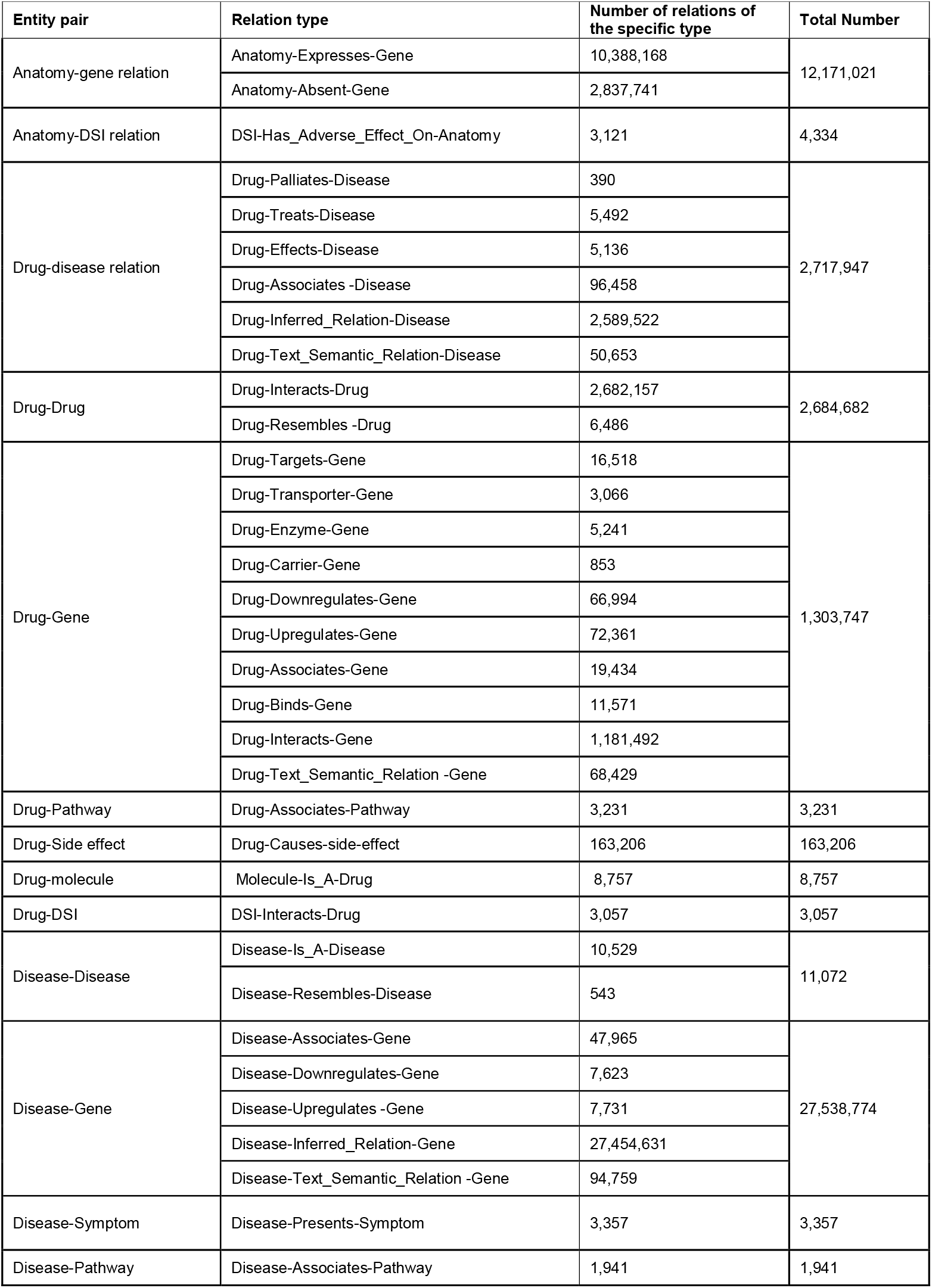
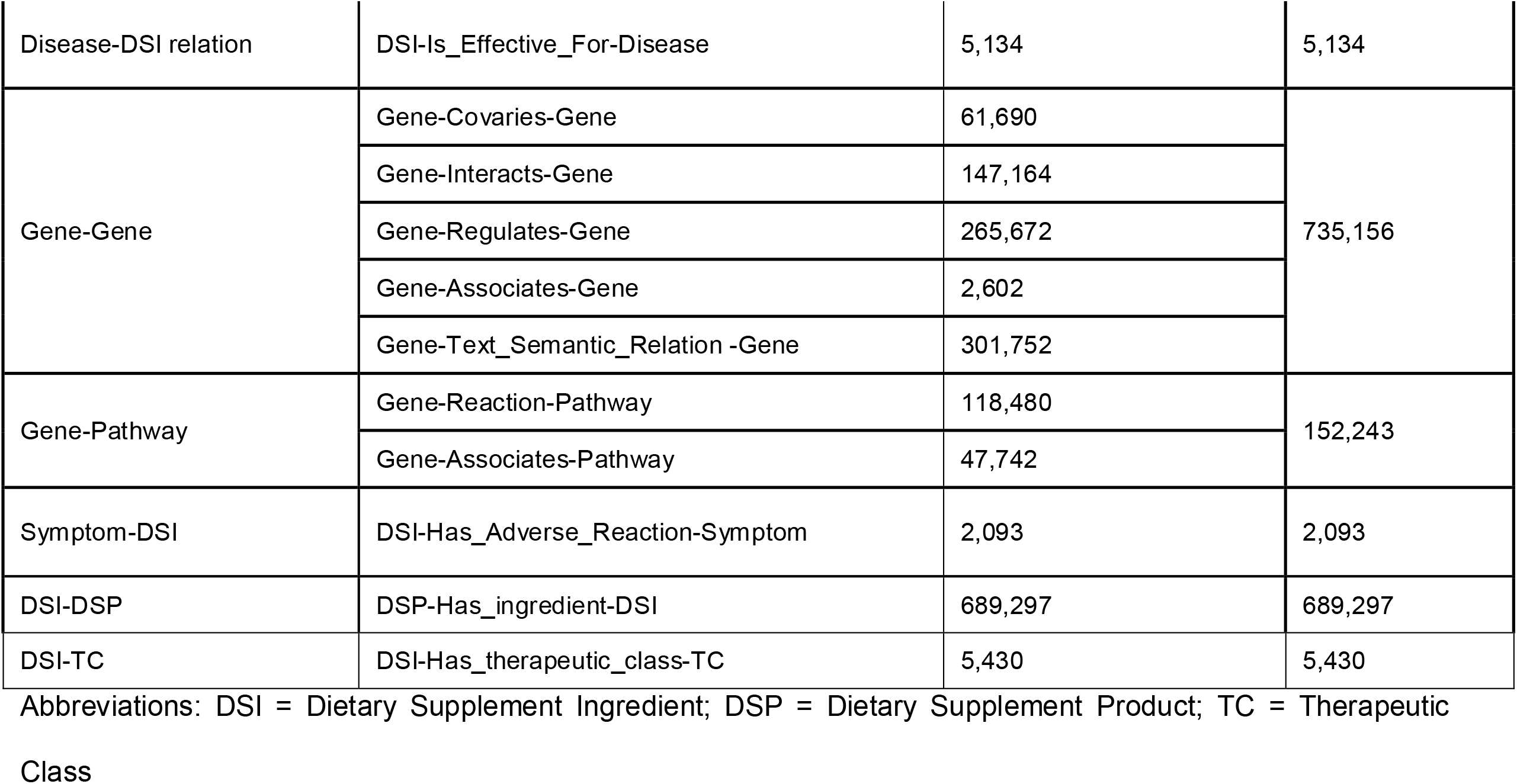
Statistics of relations among entities in iBKH.

We deployed our iBKH using Neo4j (https://neo4j.com), a robust graph database platform. We also released entity and relation source files of iBKH in CSV (comma-separated values) format, available at: https://github.com/wcm-wanglab/iBKH. Of note, the deployed version of iBKH excluded data from KEGG, as it forbids data redistribution.

### An easy-to-use interactive online portal for fast knowledge retrieval

Knowledge retrieval is the most common application scenario for a BKG like iBKH in biomedical research. In contrast to knowledge query in the traditional databases, knowledge retrieval in the iBKH needs to match the logical and structural patterns of entities and relations. This can be done by defining graph-based queries.

To fill the gap between the iBKH and biomedical and clinical researchers to facilitate its usage, we developed a web-based graphical portal that allows users to design graph-based queries for fast knowledge retrieval in a flexible, interactive manner and visualize the retrieved knowledge immediately (see **Figure 1**). Specifically, our portal has two functional modules for knowledge retrieval, i.e., biomedical entity query and path query. First, the biomedical entity query allows to retrieval information of the queried entity and its one-hop context in the iBKH, i.e., neighboring entities that directly link to the queried entity. **Figure 3a** illustrates an example of exploring biomedical context of the APOE (Apolipoprotein E) gene, which produces APOE protein and is the known major risk gene for AD (Liu et al., 2013; Strittmatter and Roses, 1995). By choosing KEGG and DrugBank in the Source section, we narrow down the query to explore entities that has relations connecting to APOE based on knowledge from the two knowledge sources. Specifically, besides AD, APOE is also associated with diseases including Sea-blue histiocytosis, Hypertriglyceridemia, Hyperlipoproteinemia type iii, and Lipoprotein glomerulopathy, which can be comorbidities of AD. APOE is associated with the AD pathway and cholesterol metabolism pathway that play a role in AD. APOE also has relations with drugs like Zinc medications (Zinc, Zinc sulfate, Zinc chloride, and Zinc acetate) that target APOE to affect progression of AD (Rivers-Auty et al., 2021; Squitti et al., 2020).

**Figure 3.**
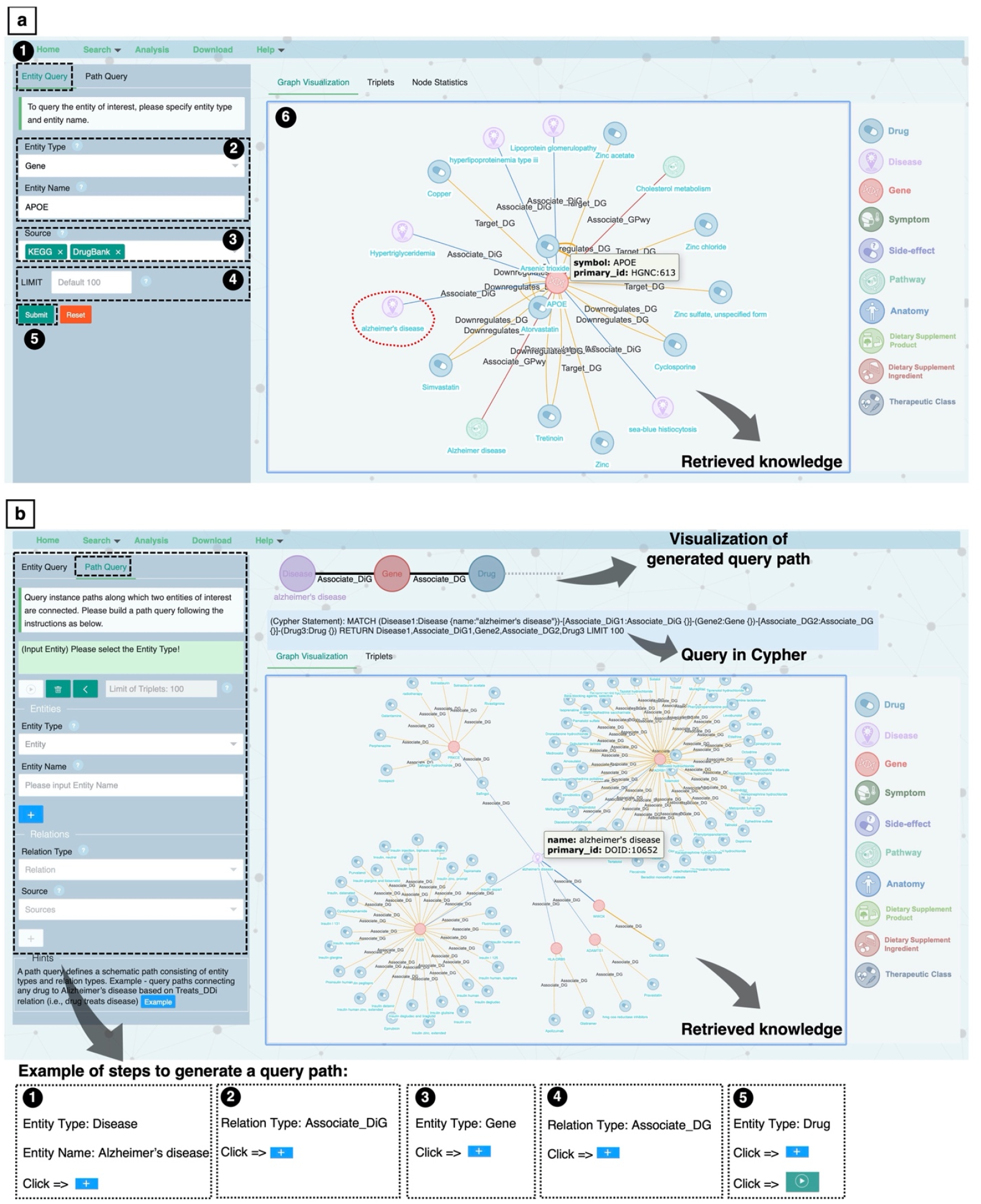
Examples of knowledge retrieval. **a**. An example of entity query – retrieving neighborhood context of APOE (Apolipoprotein E) gene in iBKH. **b**. An example of path query, retrieving drugs that connect to Alzheimer’s disease through the path *disease − [Associates_DiG] − gene − [Associats_DG] − drug*, where *Associates_DiG* and *Associats_DG* denote relation types in terms of the association between a pair of disease and gene as well as the association between a gene and a drug.

In addition, there is also a need for more sophisticated queries to retrieve multi-hop context information of the queried entity, which may help discover inconspicuous but meaningful knowledge from iBKH. **Figure 3b** illustrates an example of discovering drugs that connect to AD through the path *disease − [Associates_DiG] − gene − [Associats_DG] − drug*, where *Associates_DiG* and *Associats_DG* denote relations in terms of the “association” between a pair of disease and gene and the “association” between a pair of gene and drug, respectively. Such a query path can be generated by iteratively defining entities and relations, combined with constraints, in our portal (see **Figure 3b**). **Figure 3b** shows the retrieved knowledge. For simplicity, we only visualized 100 retrieved triplets (by setting Limit of Triplet as 100 in the portal). Centered on the AD entity, genes that have been associated with AD were first retrieved. Then, drugs that had been associated with these genes were retrieved, which can be considered as potential repurposable drugs for AD treatment. For instance, Cyclophosphamide, a medication used as chemotherapy and to suppress the immune system, is connected to the AD through the shared neighbor INSR (Insulin Receptor) gene. This is in line with previous evidence that Cyclophosphamide may help reduce cognitive decline in AD (Aisen, 2002).

### In silico hypothesis generation for Alzheimer’s disease drug repurposing

Another important application scenario for iBKH is the discovery of unknown knowledge, e.g., missing relations among entities, based on the existing, incomplete knowledge graph. In this study, we utilized a computational method for knowledge discovery in iBKH based on the advanced graph learning approaches (Nicholson and Greene, 2020; Su *et al*., 2020). As a proof of concept, we performed in silico hypothesis generation for AD drug repurposing, i.e., predicting drugs that potentially connect to the AD entity (Fang et al., 2022; Fang et al., 2021; Zeng et al., 2020; Zhou et al., 2021). We utilized knowledge graph embedding (KGE) algorithms to calculate machine-readable embedding vectors for entities and/or relations in iBKH, while preserving the graph structure (Mohamed *et al*., 2021; Su *et al*., 2020; Wang et al., 2017), using Deep Graph Library - Knowledge Embedding (DGL-KE) (Zheng *et al*., 2020). We used four advanced KGE algorithms in DGL-KE including TransE (Bordes et al., 2013), TransR (Lin et al., 2015), ComplEx (Théo et al., 2016), and DistMult (Yang et al., 2015). Then, a possibility score was calculated for each candidate drug entity based on the learned embedding vectors to measure the possibility that the drug can link to the AD via any relation(s). Such analysis has been used to identify repurposable drug candidates for COVID-19 in our previous study (Zeng *et al*., 2020). More details can be found in the **Method** section and **Figure 1**.

To evaluate performance of our method in predicting repurposable drugs for AD, we considered the FDA (Food and Drug Administration) approved drugs and drugs being tested in clinical trials for AD treatment as the ground truth, which include 10 FDA-approved, 30 in Phase IV trials, 43 in Phase III trials, 95 in Phase II trials, and 47 in Phase I trials. To avoid information leaking in prediction, all relations between the AD entity and any drug in the grand truth drug list in the iBKH were removed (see **Method** section). **Figure 4** illustrates the performance of our method. Specifically, predictions were made based on embedding vectors produced by four different KGE algorithms, i.e., TransE, TransR, ComplEx, and DistMult, respectively. We also proposed an ensemble model based on the four methods (see **Methods** section). Overall, our methods achieved desirable prediction performances, with an area under the receiver operating characteristic curve (AUC) score over 0.83 for all methods in predicting the FDA approved AD drugs, and an AUC over 0.75 in predicting FDA approved drugs and drugs in Phase IV clinical trials (n=40). In other words, the FDA approved drugs and Phase IV clinical trial drugs for AD rank higher based on our approach. In addition, the ensemble model shows a higher performance (e.g., AUC = 0.9 for FDA approved drugs, AUC = 0.79 for FDA approved plus Phase IV clinical trial drugs for AD) compared to other models, indicating that it may benefit from different KGE algorithms.

**Figure 4.**
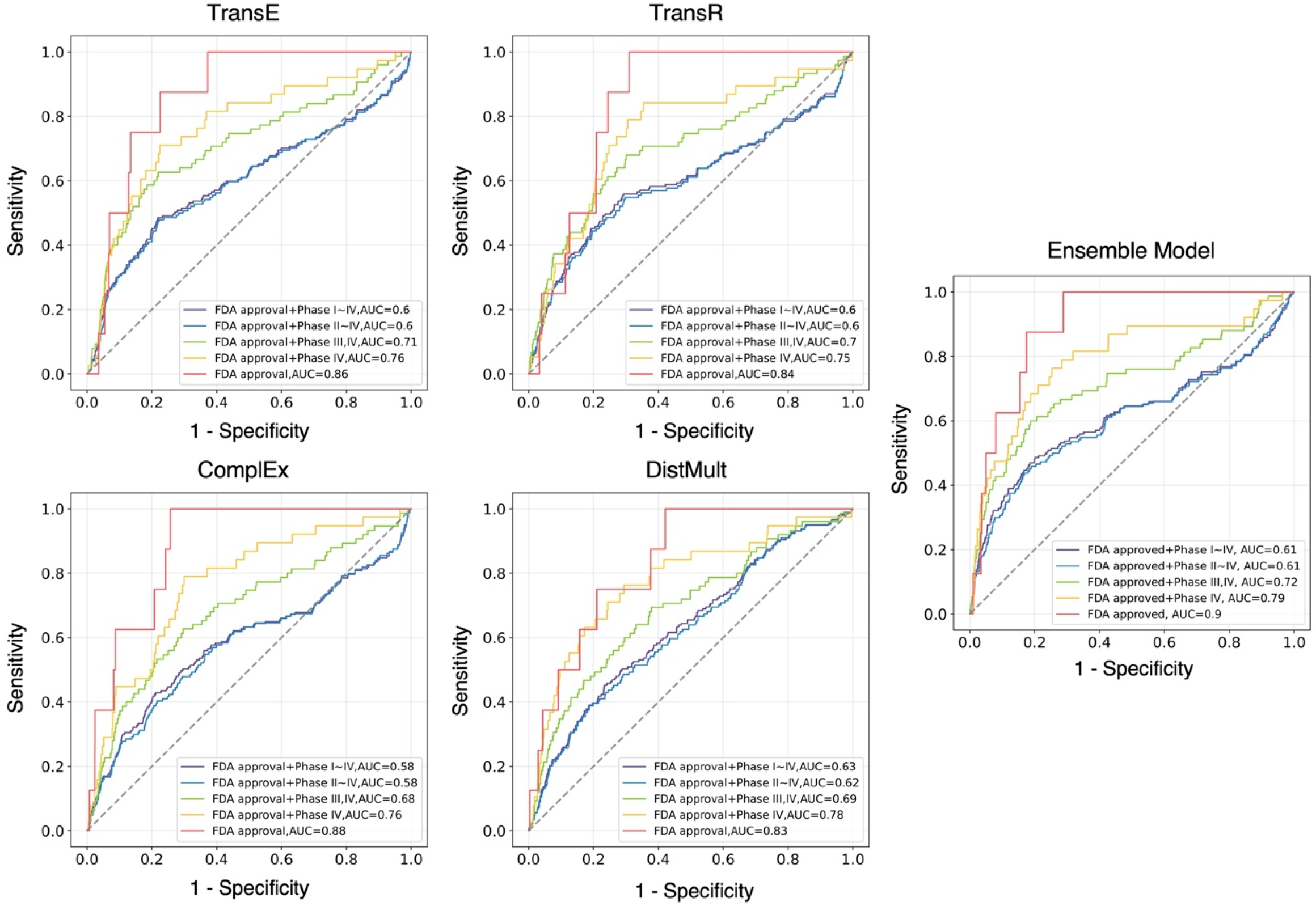
Model performance of in silicon Alzheimer’s disease drug repurposing. We used the FDA approved and clinical trial drugs for Alzheimer’s disease as ground truth. Abbreviations: AUC = area under the receiver operating characteristic curve; FDA = Food and Drug Administration.

Our model can also suggest potential drug candidates for AD, which have not been approved or involved in clinical trials for AD treatment. As a proof of concept, we highlighted the top-10 ranked potential drugs for AD treatment based on the ensemble model and iBKH (see **Table 4**).

**Table 4.** List of the top ten drugs repurposable for Alzheimer’s disease treatment.

| Rank | DrugBank ID | Drug Name | Category | Description | Notes |
| --- | --- | --- | --- | --- | --- |
| 1 | DB00836 | Loperamide | Diarrhea medication | Loperamide is used to treat diarrhea. It is often used for this purpose in inflammatory bowel disease. | Loperamide targets opioid receptors (DeHaven-Hudkins *et al.*, 1999; Giagnoni *et al.*, 1983), which has been suggested to be potentially linked to AD pathology(Cai and Ratka, 2012). |
| 2 | DB00598 | Labetalol | Anti-hypertensive drug, β-blocker | Labetalol is one of the medications called β-blockers, which is used to treat cardiovascular diseases like hypertension. | There has been evidence suggesting that β-blockers increase brain clearance of these metabolites by enhancing CSF flow. Recent studies have demonstrated that the use of β-blockers is associated with reduced risk of AD onset (Beaman *et al.*, 2022) and functional decline in AD (Rosenberg *et al.*, 2008). |
| 3 | DB00925 | Phenoxybenzamine | Anti-hypertensive drug, α-blocker | Phenoxybenzamine is an α-blocker for treating hypertension, specifically that caused by pheochromocytoma. | Phenoxybenzamine has been reported to have neuroprotective activity(Rau *et al.*, 2014). Recent drug repurposing studies have also suggested phenoxybenzamine as repurposable drug candidate to treat AD (Peng *et al.*, 2020; Williams *et al.*, 2019).. |
| 4 | DB01388 | Mibefradil | Calcium channel blocker (CCB) | Mibefradil is CCB, which was used for the treatment of hypertension and chronic angina pectoris. Mibefradil was withdrawn from the market in 1998 due to potentially harmful interactions with other drugs. | Previous studies have demonstrated that calcium dysregulation plays an important role in AD (Bojarski *et al.*, 2008). Though the usefulness of CCBs in AD remains controversial, it has shown multiple beneficial effects cell culture and animal models of AD (Anekonda and Quinn, 2011; Saravanaraman *et al.*, 2014). |
| 5 | DB00434 | Cyproheptadine | Antihistamine | Cyproheptadine is used in the treatment of allergic symptoms. | Cyproheptadine is a histamine antagonist, which has been demonstrated to reduce cognitive symptoms in AD (Zlomuzica *et al.*, 2016). |
| 6 | DB00022 | Peginterferon alfa-2b | Recombinant interferon | Peginterferon alfa-2b is used in the treatment of hepatitis B and C, genital warts, and some cancers | Peginterferon alfa-2b binds to and activates human type 1 interferon receptors, activating the JAK/STAT pathway, which has been suggested as a potential target for AD (Jain *et al.*, 2021; Nevado-Holgado *et al.*, 2019).. |
| 7 | DB00714 | Apomorphine | Dopaminergic agonist | Apomorphine is a type of dopaminergic agonist medication used for Parkinson's disease (PD) | Apomorphine is a dopamine receptor agonist for Parkinson disease and also protects against oxidative stress, which plays a role in AD (Perry *et* *et al.*, 2002). Emerging evidence showed that Apomorphine has a significant impact on improving memory function in AD (Himeno *et al.*, 2011; Nakamura *et al.*, 2017). |
| 8 | DB00623 | Fluphenazine | Antipsychotic | Fluphenazine is a phenothiazine antipsychotic medication used for treatment of psychotic disorders. | Fluphenazine is reported as a drug candidate in a recent AD drug repurposing study based on integrated network and transcriptome analysis (Peng *et al.*, 2020). |
| 9 | DB00875 | Flupentixol | Antipsychotic drug | Flupentixol is a thioxanthene neuroleptic used to treat psychotic disorders such as schizophrenia and depression. | Flupentixol is a 5-hydroxytryptamine receptor antagonist which has been reported as potential treatment for cognitive deficiency in AD (Benhamú *et al.*, 2014; Upton *et al.*, 2008). |
| 10 | DB00467 | Enoxacin | Fluoroquinolones | Enoxacin is a fluoroquinolone used for treatment of bacterial infections. | A recent study reported that appropriate use of antibiotics with macrolides and fluoroquinolones may decrease the risk of developing AD (Ou *et al.*, 2021). |

First, we found three **Anti-hypertensive drugs** ranking high based on our approach, including *<u>Labetalol</u>* (DrugBank ID: DB00598), *<u>Phenoxybenzamine</u>* (DrugBank ID: DB00925), and *<u>Mibefradil</u>* (DrugBank ID: DB01388). Specifically, Labetalol is a type of *β*-blockers. There has been evidence suggesting that *β*-blockers increase brain clearance of these metabolites by enhancing cerebrospinal fluid flow. Recent studies have demonstrated that the use of *β* - blockers is associated with reduced risk of AD onset (Beaman et al., 2022) and functional decline in AD (Rosenberg et al., 2008). Phenoxybenzamine is an *a*-blocker, which has been reported to have neuroprotective activity (Rau et al., 2014). Recent drug repurposing studies have also suggested phenoxybenzamine as a repurposable drug candidate to treat AD (Peng et al., 2020; Williams et al., 2019). Mibefradil is a calcium channel blocker (CCB). Though Mibefradil was withdrawn from the market in 1998 due to harmful interactions with other drugs, our findings may suggest CCB as potential candidate for AD because that calcium dysregulation has been reported to play a role in AD (Bojarski et al., 2008) and CCB has shown multiple beneficial effects cell culture and animal models of AD (Anekonda and Quinn, 2011; Saravanaraman et al., 2014).

Second, we found two **Antipsychotic drugs** as candidates for AD treatment: *<u>Fluphenazine</u>* (DrugBank ID: DB00623) and *<u>Flupentixol</u>* (DrugBank ID: DB00875). Fluphenazine has been reported as a drug candidate in a recent AD drug repurposing study based on integrated network and transcriptome analysis (Zhao et al., 2020b). On the other hand, Flupentixol is a 5-hydroxytryptamine receptor antagonist, which has been reported as potential treatment for cognitive deficiency in AD (Benhamú et al., 2014; Upton et al., 2008).

We also found other candidate drugs for AD, including *<u>Loperamide</u>* (DrugBank ID: DB00836), *<u>Cyproheptadine</u>* (DrugBank ID: DB00434), *<u>Peginterferon alfa-2b</u>* (DrugBank ID: DB00022), *<u>Apomorphine</u>* (DrugBank ID: DB00714), and *<u>Enoxacin</u>* (DrugBank ID: DB00467). In particular, Loperamide is used to treat diarrhea. Previous studies reported that Loperamide targets opioid receptors (DeHaven-Hudkins et al., 1999; Giagnoni et al., 1983), which has been suggested to be potentially linked to AD pathology(Cai and Ratka, 2012). Cyproheptadine belongs to the histamine antagonists, which have been demonstrated to reduce cognitive symptoms in AD (Zlomuzica et al., 2016). Peginterferon alfa-2b is a recombinant interferon, which is used in the treatment of hepatitis B and C, genital warts, and some cancers. Peginterferon alfa-2b has been reported to bind to and activate human type 1 interferon receptors. Such a procedure activates the JAK/STAT pathway, which has been suggested as a potential target for AD (Jain et al., 2021; Nevado-Holgado et al., 2019). Apomorphine is a dopamine receptor agonist for Parkinson’s disease (PD). It can protect against oxidative stress, which plays a role in AD pathology (Perry et al., 2002). Emerging evidence showed that Apomorphine has a significant impact on improving memory function in AD (Himeno et al., 2011; Nakamura et al., 2017). Enoxacin belongs to the fluoroquinolones, which is used for treatment of bacterial infections. A recent study reported that appropriate use of antibiotics with macrolides and fluoroquinolones may decrease the risk of developing AD (Ou et al., 2021).

## Discussions

Due to the unparalleled development rate of novel techniques in biomedical research and healthcare, a massive and continuously increasing volume of biomedical knowledge has been produced in the past decades. In addition, the biomedical knowledge captured from different sub-domains of biomedicine is typically stored in different types of databases. These include biomedical ontologies (Rubin *et al*., 2008; Smith *et al*., 2005) that provide hierarchical relationship-based descriptions for entities from a specific entity type and manually curated knowledge bases that focuses on a specific sub-domain in biomedicine (Callahan *et al*., 2020; Zhu *et al*., 2019). To better use the rich biomedical knowledge while overcoming the massive volume and data heterogeneity, there is the need for harmonizing and integrating the biomedical knowledge from data sources across diverse sub-domains in biomedicine. To this end, we curated a comprehensive BKG, the iBKH, through collecting, managing, and cleaning raw data from diverse sources, creating standardized vocabularies for biomedical entity normalization, as well as knowledge integration. To date, our iBKH has collected biomedical knowledge from 18 sources, including not only the biomedical ontologies and biomedical knowledge bases, based on which most existing BKGs have been built, but also existing BKGs that have integrated diverse sources. In addition to the general entity types that are commonly studied in biomedicine, such as genes, diseases, drugs, pathways, etc., our iBKH also involves our previously curated dietary supplement knowledge base, the iDISK (Rizvi *et al*., 2020). Research studies have demonstrated that the dietary supplements play a role in human diseases, such as AD (Luchsinger and Mayeux, 2004; Luchsinger et al., 2007), cancers (Williams and Hord, 2005), diabetes (van Dam et al., 2002), etc. We believe that involvement of the dietary supplement knowledge will provide complementary knowledge for better human healthcare.

We deployed the iBKH publicly available in both tabular format (CSV files) and Neo4j, which allows fast knowledge retrieval through creating Cypher queries. Though Cypher, inspired by the SQL, is relatively easy to learn, there remains a gap between the iBKH in Neo4j and biomedical researchers and healthcare providers, as knowledge retrieval in a graph needs to define the queries by matching the logical and structural patterns of entities and relations, which are more complex than the SQL queries. As a result, we developed a web-based graphical portal, which allows users to design the desired graph query using a graphical UI. The query is translated to Cypher query in the back end for fast knowledge retrieval and the retrieved knowledge (typically a sub-graph from the iBKH) is visualized in the portal immediately. In this way, the iBKH is more user-friendly, providing users who have no Cypher programming experience the large flexibility to implement efficient biomedical knowledge retrieval.

In addition, we implemented the advanced graph learning approaches to the iBKH for novel biomedical knowledge discovery. The graph learning approaches is a branch of machine learning and artificial intelligence (AI), which devote to building learning algorithms to model graph structure for discovering unobserved knowledge. In this study, we utilized the DGL-KE (Deep Graph Library - Knowledge Embedding) (Zheng *et al*., 2020), a Python-based implementation for advanced knowledge graph embedding algorithms, for efficient and scalable graph learning in iBKH. As a proof of concept, we demonstrated a use case of iBKH, armed with the graph learning algorithms, for in silico hypothesis generation for AD drug repurposing. We not only observed good performance of our method, using the FDA-approved drugs and clinical trial drugs for AD as ground truth, but also identified repurposable drugs for AD treatment. By manual literature review, we found evidence supporting potential of the identified candidate drugs to treat AD. More importantly, our iBKH-based knowledge discovery pipeline is flexible and feasible, and can be applied to more diseases of interest beyond AD, by predicting potential relations between the disease of interest and drugs in iBKH. Our pipeline can also adapt to other biomedical application scenarios, such as prioritizing risk genes of disease (gene-disease relation prediction), predicting candidate target protein for drugs (drug-gene relation prediction), identifying potential drug-drug interactions (drug-drug relation prediction), etc.

### Limitations of the study

Our iBKH has a few limitations. First, the procedures of constructing and curating iBKH rely on sophisticated efforts of raw data file extraction and pre-processing, data annotation, as well as terminology normalization, which may lead to **incorrectness**, referring to facts in the iBKH that is inconsistent with real-world evidence. To address this, we utilized the well-designed biomedical vocabularies such as the Unified Medical Language System (UMLS) to enhance entity term normalization, which can help reduce the risk of errors caused by the ambiguous biomedical entities. We also performed manual review to reduce incorrectness. More specifically, the integrated file for each entity type or relation type underwent multiple rounds of manual review based on random sampling with replacement. Even so, due to the massive volume of iBKH, there remains the need for a more efficient way to address incorrect facts in iBKH. Graph learning algorithms for knowledge graph refinement is a potential solution in this context. For instance, our early effort in graph learning-based knowledge graph refinement could be extended to address this issue (Zhao *et al*., 2020b).

Another issue is knowledge **incompleteness**. We built the iBKH by collecting and integrating data from diverse sources. It includes knowledge in a board range of sub-domains of human health. However, incompleteness is still inevitable. On one hand, abundant knowledge sources have been online available and there are good sources that are not involved in iBKH yet. On the other hand, there remains massive biomedical knowledge that has not yet been discovered or is deep buried in the noisy biomedical and health data and literature. In this context, some studies have been focused on deriving knowledge from biomedical literature (Xu et al., 2013; Zhang et al., 2018; Zhao et al., 2021) or human healthcare data such as the EHR (electronic health records) (Chen et al., 2019; Rotmensch *et al*., 2017). The derived knowledge could be a good complement for our iBKH. In addition, the use of graph learning algorithm to discover hidden knowledge based on the existing iBKH graph structure is another solution and needs more attention in our future work.

Like most existing BKGs, e.g., Hetionet and CKG, our iBKH focuses on the general biomedical knowledge. For the sake of precision medicine on some specific human diseases or health conditions, there is the need for more fine-grained knowledge with a specific focus on them. For instance, COVID-KG (Wang et al., 2020) included biomedical knowledge with a specific focus on COVID-19; KGHC (Li *et al*., 2020) is a knowledge graph constructed focusing on addressing hepatocellular carcinoma. Following this idea, we will adapt our iBKH to address problems in specific diseases and health conditions like AD, Parkinson’s disease, and mental illness. For example, we plan to collect the fine-grained data, such as genotype-phenotype associations and brain region atrophy-phenotype associations and incorporate them to enrich iBKH, for the specific usage of these diseases.

Last, there is the need of further validation for the discovered novel knowledge from iBKH. To this end, our future work will also focus on knowledge validation by leveraging advanced data science techniques. On one hand, we plan to build a knowledge validation system based on biomedical text mining (Zhao *et al*., 2021). For instance, leveraging our previous biomedical literature retrieval/matching algorithms (Zhao et al., 2020a; Zhao et al., 2019), we will be able to identify evidence from massive biomedical literature resource, supporting the identified novel triplets (i.e., knowledge) in iBKH. We plan to add this new functionality to iBKH portal. On the other hand, for drug repurposing hypothesis generation, we will validate treatment efficiency of the identified repurposable drug candidates for target disease, such as AD, using our computational clinical trial emulation approach (Zang et al., 2022) based on real-world clinical data.

## Data Availability

The data is currently available from the corresponding author on reasonable request.

## Author Contributions

FW for conceptualization, investigation, writing, reviewing, and editing of the manuscript. CS for investigation, drafting, editing, and reviewing manuscript. YH led the effort on data preparation, knowledge graph construction, data analysis and web interface implementation. SR, JM, ZA for improving data standardization and organization, efficiency of user interface, and language of the manuscript. HZ, FFC, and GG for data collection and data preparation. AC for knowledge graph embedding implementation. ZB for critical discussion on constructing iBKH. WG for knowledge graph quality check. JT and YL for critical discussion on knowledge graph embedding algorithms. FC, RZ and JB for discussion, design, and interpretation of the case study on AD. All authors have given approval to the final version of the manuscript.

## STAR*METHODS

### Resource Availability

#### Lead contact

Further information should be directed to and will be fulfilled by the lead contact, Dr. Fei Wang

#### Materials Availability

- The harmonized entity and relation source files for iBKH in CSV (comma-separated values) format are publicly available online at https://github.com/wcm-wanglab/iBKH/tree/main/iBKH.
- The iBKH online portal is publicly available at http://ibkh.ai/.

The deployed version of iBKH excluded data from KEGG, as it forbids data redistribution.

### Data and Code Availability

- This paper integrates publicly available biomedical knowledge bases. These accession URLs for the knowledge bases are listed in the key resources table.
- The computer codes for iBKH construction and iBKH-based knowledge discovery are publicly available online at https://github.com/wcm-wanglab/iBKH/tree/main/Codes.
- Any additional information required to reanalyze the data reported in this paper is available from the lead contact upon request.

### METHOD DETAILS

#### Overview

Our ultimate goal was to build a biomedical knowledge graph via comprehensively incorporating biomedical knowledge as much as possible. To date, we have collected and integrated 18 publicly available data sources, harmonized and consolidated them into a comprehensive data compendium. Details of the used data sources were listed in **Table 1**.

#### Raw data processing

Given the data sources, the first step was to pre-process the raw files of them and extract knowledge, including entity information and relation information. Generally, the databases release their raw data files in various formats, such as comma-separated values (CSV), tab-separated values (TSV), TXT, EXCEL tablet, Hypertext Markup Language (HTML), Resource Description Framework (RDF), and Web Ontology Language (OWL). To address this, for each database, we parsed the raw files and extracted structured data, i.e., the descriptive files for each type of biomedical entity and the files of each type of relation. Such procedure varies by databases or even by files within the same database.

#### Term harmonization

To integrate data from diverse sources, there is a need for harmonizing the entity terms. To achieve this, we utilized a greedy strategy. Specifically, for a specific entity type, we first chose a database to initialize the entity vocabulary. Next, we built a linkage pool, containing multiple identifiers of the given entity type, to map and integrate entities from all databases to enrich the entity vocabulary one by one.

For **gene** entity type, we used the HUGO Gene Nomenclature Committee (HGNC) gene repository (Braschi *et al*., 2019) as the initial vocabulary of gene entities, as it defines a standard nomenclature for human the genes. The linkage pool for normalization included HGNC IDs, HGNC symbols, and National Center for Biotechnology Information (NCBI) IDs.

For **drug** entity type, we initialized our vocabulary using DrugBank (Wishart *et al*., 2018) as it provides the up-to-date list of approved drugs and investigational drugs under clinical trials. The linkage pool for drug entity normalization included DrubBank IDs, Medical Subject Heading (MeSH) terms, MeSH term IDs, Unified Medical Language System (UMLS)(Bodenreider, 2004) Concept Unique Identifiers (CUIs), and the drug names in UMLS.

For **molecule** entity type, we used the ChEMBL(Gaulton *et al*., 2012), a manually curated database of molecules with drug properties, for initializing the vocabulary. The linkage pool for the molecule entities normalization included ChEMBL IDs and International Chemical Identifier (InChi).

For **Side-Effect** entity type, we collected the side-effect entities from the SIDER(Kuhn *et al*., 2016) and described them by using the UMLS CUIs.

For **disease** entity type, we used the Disease Ontology (Schriml *et al*., 2012) for initializing the vocabulary, as it is a structured database of diseases based on etiological classification. The linkage pool we used for the disease entity normalization included Disease Ontology IDs, MeSH terms, MeSH term IDs, UMLS CUIs, and the disease names in UMLS.

For **symptom** entity type, we collected the symptom entities from the Hetionet (Himmelstein *et al*., 2017) and integrated Dietary Supplements Knowledge (iDISK) (Rizvi *et al*., 2020), and described them by using the MeSH term and MeSH term ID. We used UMLS CUI as the linkage for symptom entities normalization.

For **Pathway** entity type, we used the Reactome (Fabregat *et al*., 2018), a manually curated and peer-reviewed pathway database, for initializing the vocabulary. The linkage pool for the pathway entities normalization contained the Reactome IDs, Gene Ontology IDs, and KEGG IDs.

For **anatomy** entity type, we used the Uberon (Mungall *et al*., 2012) for initializing the vocabulary, as it is a cross-species anatomical ontology based on traditional anatomical classification. The linkage pool for the anatomy entities harmonization included Uberon IDs, MeSH terms, MeSH term IDs, UMLS CUIs, and the anatomy names in UMLS.

For **Dietary Supplement Ingredient (DSI), Dietary Supplement Product (DSP)**, and **Therapeutic Class (TC)** entities, data were collected from our previous curated iDISK (integrated Dietary Supplements Knowledge) (Rizvi *et al*., 2020). We used iDISK concept IDs and UMLS CUIs (for TCs) to describe them.

#### Knowledge integration

After the above normalization procedures, we obtained a CSV file for each entity type, storing all normalized entity terms of the specific entity type followed by their synonyms and detailed descriptions. We were then able to integrate knowledge extracted from different knowledge bases to build iBKH. Specifically, in a BKG, a basic knowledge unit is a triplet, typically defined as *<****head entity, relation, tail entity****>*, which indicates that there exists a relation from the ***head entity*** to the ***tail entity*** in iBKH. Of note, for each pair of head entity and tail entity, there can be multiple types of relations. For instance, we stored “targets”, “Transporter”, “Enzyme”, “Carrier”, “downregulates”, “upregulates”, “associates”, “binds”, “interacts”, and “text_semantic” relations between drugs and genes. We also stored the data source information, indicating from which data source(s) we acquired the specific triplet.

#### iBKH deployment based on graph database

We deployed our curated BKG, i.e., the iBKH, using Neo4j (https://neo4j.com), a well-designed graph database platform that allows structured queries in a grap. Specifically, Neo4j can take the CSV files of entities and relations we generated above as input and automatically created a KG instance. In this way, the iBKH can be updated efficiently and flexibly.

#### Graphical portal for fast knowledge retrieval

We developed a web-based graphical portal, which allows the users to design graph query paths visually and flexibly and translates them into Cypher queries (query language provided by Neo4j) automatically in the back end. Specifically, we built the back end (i.e., the server side) using Django (https://www.djangoproject.com/), a high-level Python-based web framework. The iBKH, stored in Neo4j, was linked to the back end. The front end (i.e., the web application side) was built based on HyperText Markup Language Version 5 (HTML5), and Cascading Style Sheets (CSS). JavaScript-based software, the neovis (https://github.com/neo4j-contrib/neovis.js/) and D3.js (https://d3js.org/), were used for graph visualization and data exploration and visualization, respectively.

#### iBKH-based knowledge discovery

We developed a machine learning pipeline for knowledge discovery in the iBKH, which contains two steps as follows.

##### Step 1, knowledge graph embedding (KGE) learning

The goal of KGE is to learn embeddings, i.e., meaningful and machine-readable vector-based representations for entities and/or relations in iBKH, while preserve the graph structure (Goyal and Ferrara, 2018; Su *et al*., 2020; Wang *et al*., 2017). In biomedicine, the learned embeddings (i.e., vector representations) of biomedical entities and relations can be used in accelerating diverse down-stream research tasks, such as drug implication discovery (Nicholson and Greene, 2020; Zhang *et al*., 2021; Zheng et al., 2021; Zhu *et al*., 2020), multi-omics data analysis (Nicholson and Greene, 2020; Santos *et al*., 2022), clinical data (e.g., electronic healthcare record) analysis (Choi et al., 2017; Nelson *et al*., 2019), and knowledge extraction from biomedical literature (Wang *et al*., 2020). In this work, we used the Deep Graph Library - Knowledge Embedding (DGL-KE) (https://github.com/awslabs/dgl-ke)(Zheng *et al*., 2020), a Python-based implementation for the advanced KGE algorithms, such as TransE (Bordes *et al*., 2013), TransR (Lin *et al*., 2015), ComplEx (Théo *et al*., 2016), and DistMult (Yang *et al*., 2015). Using the advanced multi-processing and multi-GPU (graphics processor unit) techniques, the DGL-KE accelerates the learning procedures in large-scale graphs like iBKH.

##### Step 2, link prediction

The task can be formulated as predicting the probability that an unobserved triplet < *h, r, t* > exists in the iBKH, where *h* and *t* are the head and tail entities, and *r* is the potential relation, respectively. Specifically, we defined a possibility score of a candidate triplet < *h, r,t* > as *PS*(< *h, r, t* >) = *sigmoid*([(*h, r, t*)). The sigmoid function is defined as *sigmoid*(*a*) = 1/(1 exp (-*a*)). *f*(·) is the scoring function of the KGE algorithm we used to calculate the embedding vectors.

- TransE, *f*(*h, r, t*) = - ∥***h + r - t***∥_*p*_, where ***h, r, t*** are the embedding vectors of *h, r, t*, respectively.
- TransR, 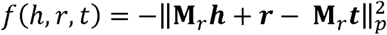, where **M**_*r*_ is a projection matrix for each relation *r* that project entities *h* and *t* to semantic space of the relation.
- ComplEx, *f*(*h, r, t*) = <*Re*(***h***), *Re*(***r***), *Re*(***t***)> + <*Im*(***h***), *Im*(***r***), *Im*(***t***)> + <*Re*(***h***), *Im*(***r***), *Im*(***t***)> − <*Im*(***h***), *Im*(***r***), *Re*(***t***)>, where *Re*(***x***) and *Im*(***x***) are the real and imaginary parts of the complex valued vector *x*, respectively.
- DistMult, *f*(*h, r, t*) = ***h***^*T*^**W**_*r*_ ***t***^*T*^, where **W**_*r*_ is relation matrix, which is restricted to a diagonal matrix.

Summarized details of the KGE algorithms can be found elsewhere (https://dglke.dgl.ai/doc/kg.html).

##### In silico hypothesis generation for Alzheimer’s disease drug repurposing

As a proof of concept, we performed in silico hypothesis generation for Alzheimer’s disease (AD) drug repurposing, which is to predict potential drug entities that can be linked to the AD entity with a ‘treats’ relation in the iBKH. To this end, we first downloaded all Food and Drug Administration (FDA) approved drugs and drugs in clinical trials (Phases I-IV) for AD from the DrugBank (https://go.drugbank.com/), constructing the grand truth drug list. Specifically, we obtained a total of 10 FDA-approved drugs, 30 drugs in Phase IV trials, 43 drugs in Phase III trials, 95 drugs in Phase II trials, and 47 drugs in Phase I trials for AD treatment. Next, to avoid information leaking in prediction, all relations between the AD entity and any drug in the grand truth drug list in the iBKH were removed. Then, entity and relation embedding vectors were calculated using the KGE algorithms. After that, we calculated possibility scores for potential all < *e*_*d*_, *r, e*_*D*_ > triplets, where *e*_*d*_ indicates any drug entity, *e*_*D*_ indicates the AD entity, and *r* indicates a relation between them. The drugs were ranked based on the possibility scores. In this study, we calculated the possibility scores based on four KGE algorithms, i.e., TransE (Bordes *et al*., 2013), TransR (Lin *et al*., 2015), ComplEx (Théo *et al*., 2016), and DistMult (Yang *et al*., 2015). To enhance prediction, we also proposed an ensemble model. Specifically, the rank of drug *e*_*d*_ in the ensemble model was defined as *PS*^*ensemble*^(< *e*_*d*_, *r, e*_*D*_ >) = ∑_*i*_(*N*^*Dr*^ − *Rank*^*i*^(< *e*_*d*_, *r, e*_*D*_ >)) where *i* indicates the *i*-th KGE algorithm and *N*^*Dr*^ indicates total number of drugs in iBKH.

To evaluate prediction performance, we compared the top *K* ranked drugs with the ground truth drugs. By sliding the value of *K*, we were able to produce the receiver operating characteristic curve (ROC) and the area under ROC (AUC) score.

Finally, we re-trained the KGE models without removing known relations between AD and drug entities and used the embeddings to predict novel repurposable drug candidates for AD treatment. For the predicted drugs that potentially link to AD, we performed manual literature review to identify supporting evidence of the prediction.

